# Polygenic modelling of treatment effect heterogeneity

**DOI:** 10.1101/2020.01.06.20016618

**Authors:** Zhi Ming Xu, Stephen Burgess

## Abstract

Mendelian randomization is the use of genetic variants to assess the effect of intervening on a risk factor using observational data. We consider the scenario in which there is a pharmacomimetic (that is, treatment-mimicking) genetic variant that can be used as a proxy for a particular pharmacological treatment that changes the level of the risk factor. If the association of the pharmacomimetic genetic variant with the risk factor is stronger in one subgroup of the population, then we may expect the effect of the treatment to be stronger in that subgroup. We test for gene–gene interactions in the associations of variants with a modifiable risk factor, where one genetic variant is treated as pharmacomimetic and the other as an effect modifier, to find genetic sub-groups of the population with different predicted response to treatment. If individual genetic variants that are strong effect modifiers cannot be found, moderating variants can be combined using a random forest of interaction trees method into a polygenic response score, analogous to a polygenic risk score for risk prediction. We illustrate the application of the method to investigate effect heterogeneity in the effect of statins on low-density lipoprotein cholesterol.

## Introduction

Genetic variants can be treated as proxies for treatments to assess the effect of intervening on a particular biological pathway using observational data [Thanassoulis and O’Donnell, 2009; Plenge et al., 2013]. For example, 3-hydroxy-3-methylglutaryl–coenzyme A reductase (HMGCR) inhibitors (known as statins) have been developed to reduce levels of low-density lipoprotein (LDL) cholesterol. Variants in the *HMGCR* gene region that predispose individuals to having higher or lower average levels of LDL-cholesterol can be used to predict the effect of HMGCR inhibitors on disease outcomes [Khera and Rader, 2009]. Associations between the *HMGCR* variants and coronary artery disease risk suggest that statins should reduce coronary artery disease risk [Ference et al., 2015], as has been observed in clinical trials [Cholesterol Treatment Trialists’ Collaboration, 2005]. The approach of using genetic variants to make causal inferences from observational data is known as Mendelian randomization [Davey Smith and Ebrahim, 2003; Burgess and Thompson, 2015].

An extension of Mendelian randomization known as ‘factorial Mendelian randomization’ uses genetic variants in two gene regions to assess treatment interactions [Rees et al., 2019]. For example, genetic variants in the *HMGCR* gene region, representing proxies for statins, and genetic variants in the proprotein convertase subtilisin–kexin type 9 (*PCSK9*) gene region, representing proxies for PCSK9 inhibitors, showed no interaction in their associations with either LDL-cholesterol or coronary artery disease [Ference et al., 2016]. This suggests statins should lower LDL-cholesterol when used in conjunction with PCSK9 inhibitors, with no dilution in their effect. Alternatively, we can consider the interaction between an *HMGCR* variant and a genetic variant in a gene region that does not correspond to a pharmacological intervention, but instead is viewed as a stratifying variable. In this case, a statistical interaction would be interpreted as effect modification – the association of the *HMGCR* variant (and, by inference, the effect of statins) differs for individuals in different genetic subgroups defined by the stratifying variable [VanderWeele, 2015]. Effect modification has implications for precision medicine, as individuals for whom statins are more effective could be prescribed statins earlier in life. Henceforth, when considering gene–gene interactions, we interpret variants in one gene region as proxies for the treatment (referred to as pharmacomimetic variants) and other variants as stratifying variables (referred to as moderating variants).

Few interactions between genetic variants have been robustly demonstrated [Cordell, 2009]. There are several possible reasons for this: in particular, i) genetic variants typically have small effects on phenotypes and interaction effects are generally smaller in magnitude than main effects; and ii) hypothesis testing approaches must incorporate correction for multiple testing, which lowers power to detect a true interaction. In the context of risk prediction, polygenic risk scores have been used successfully to overcome the problems of small effects and multiple testing. By summing the contributions of large numbers of variants across the whole genome into a single univariable score, prediction is improved compared to approaches that take information on a small number of variants [Dudbridge, 2013; Inouye et al., 2018]. This suggests the possibility of using a similar approach to construct genetic subgroups of the population which differ in their predicted response to pharmacological treatment, even if no individual variants can be found that have a strong gene–gene interaction.

In this paper, we introduce an agnostic approach to create a composite polygenic response score that predicts treatment effect heterogeneity similarly to a polygenic risk score for risk prediction. As a proof-of-concept example, we demonstrate the approach for the effect of statins on LDL-cholesterol. We first construct a pharmacomimetic score for statins from genetic variants proximal to the *HMGCR* gene region which have been shown to be associated with LDL-cholesterol [Ference et al., 2016]. We then perform a genome-wide search for moderating variants in the training subset of the data, and combine these variants using the random forest of interaction trees method [Su et al., 2018, 2009]. We proceed to verify in the validation subset whether the predicted treatment effects for different genetic subgroups are more variable than expected solely based on chance, and consider the impact of leaf node sizes and interaction significance thresholds on the variability of estimates. We conclude by discussing the applicability of this approach to wider practice in the emerging area of precision medicine.

Software for implementing the method is available from https://github.com/zmx21/polyresponse.

## Methods

### Set-up and notation

We consider a risk factor *x*_*i*_ for individuals *i* = 1, …, *N*, a pharmacomimetic genetic variant *z*_*i*_ which can be considered as a proxy for a particular intervention on the risk factor, *J* candidate moderating variants *g*_*ij*_ where *j* = 1, …, *J*, and *K* measured covariates *c*_*ik*_ where *k* = 1, …, *K*. The pharmacomimetic genetic variant does not have to be one single nucleotide polymorphism (SNP), but could instead be a weighted score, representing the predicted values of the risk factor based on genetic variants in the pharmacomimetic gene region. This would be a worthwhile strategy if there were multiple variants independently associated with the risk factor in that gene region. We consider the following linear regression model:

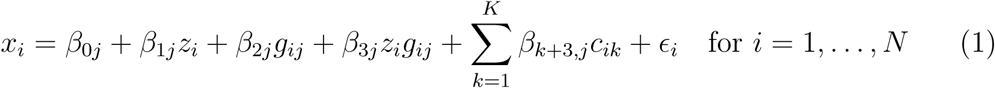

The main parameter of interest is *β*_3*j*_, representing the interaction between the pharmacomimetic genetic variant and the *j*th moderating variant. The marginal association between the pharmacomimetic genetic variant and the risk factor is *β*_1*j*_ + *β*_3*j*_*g*_*ij*_. If *β*_3*j*_ = 0, then the association between the pharmacomimetic genetic variant and the risk factor does not depend on the value of *g*_*ij*_. Whereas if *β*_3*j*_ ≠ 0, then the association between the pharmacomimetic genetic variant and the risk factor is stronger for some values of *g*_*ij*_.

If we find a genetic variant with statistically robust evidence for an interaction, then we can use this variant to divide the population into genetic groups which differ in their expected response to the treatment. For example, if the marginal association between the pharmacomimetic variant and the risk factor is zero for individuals with *g*_*ij*_ = 0, but positive for individuals with *g*_*ij*_ > 0, this suggests that the corresponding pharmacological intervention on the risk factor is likely to only influence the risk factor in individuals with *g*_*ij*_ > 0, and have no average effect in those with *g*_*ij*_ = 0. However, it is unlikely that there are many individual variants with strong interactions. This motivates the development of approaches for combining variants that display some evidence of interaction into a composite genetic moderator.

### Interaction tree

The random forest of interaction trees method is a recursive partitioning approach that was introduced by Su et al. [2009]. We first present how to construct a single interaction tree, and then how to construct a random forest of interaction trees. For each candidate split variant *G*_*j*_, we consider an indicator variable *I*_*i*_ for individuals indexed by *i* in two ways: i) *I*_*i*_ = 0 for the subgroup with *g*_*ij*_ = 0 versus *I*_*i*_ = 1 for the subgroup with *g*_*ij*_ = 1, 2; and ii) *I*_*i*_ = 0 for *g*_*ij*_ = 0, 1 versus *I*_*i*_ = 1 for *g*_*ij*_ = 2. We then calculate the t-statistic for the interaction term *γ*_3_ from the model:

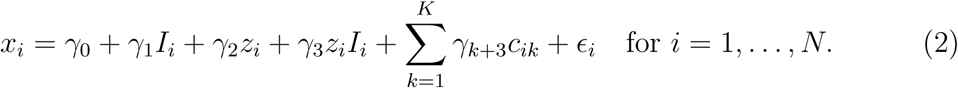

We split the sample into two subgroups based on the candidate variant and indicator variable combination with the greatest squared t-statistic. We continue recursively to split each subgroup in the same way until an additional split results in a daughter node which is below a threshold for minimum node size, at which point a terminal node is created without the additional split. The association of the pharmacomimetic variant with the risk factor (the treatment effect) is calculated for each terminal node. An example tree is shown in Figure 1.

**Figure 1:**
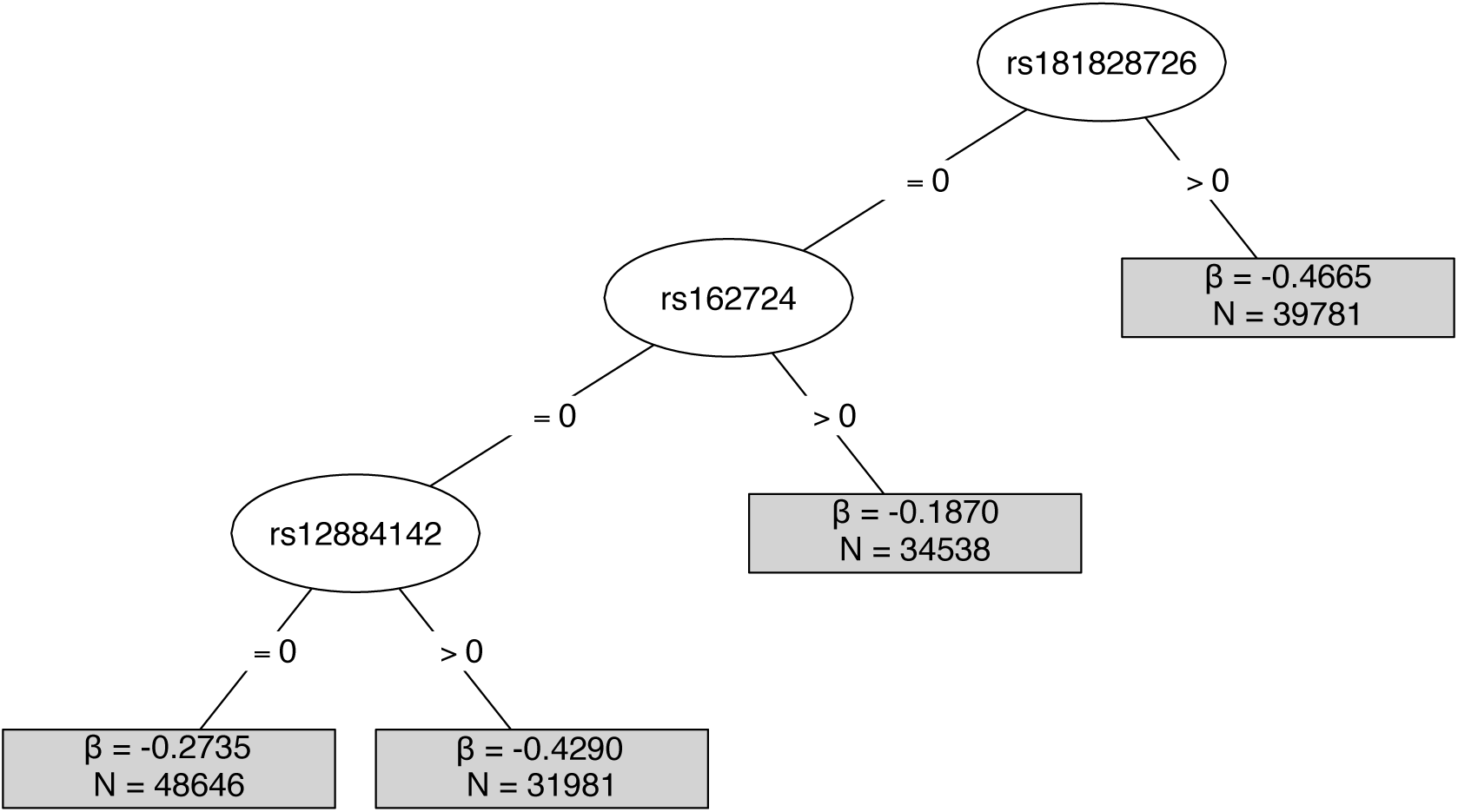
Example of a single interaction tree, constructed with interaction significance threshold *p* < 7 × 10^−6^ and minimum node size of 30 000. Terminal nodes show the predicted treatment effect estimate for that subgroup (*β*) and the size of the subgroup (N).

### Random forest of interaction trees method

The random forest is an ensemble method which aims to reduce variance (overfitting) inherent in individual interaction trees by aggregating multiple trees constructed from bootstrap samples [James, 2013; Breiman, 2001]. To construct a random forest of interaction trees, we initially split the dataset at random into a training set (2*/*3 of the sample) and a validation set (1*/*3 of the sample). We take 2000 bootstrap samples of 2*/*3 of the training set (4*/*9 of the total dataset) and construct an interaction tree for each sample. A random set of candidate split variants (3*/*4 of all available moderating variants) is considered at each search for an optimal interaction term to decorrelate the trees within the random forest and so further reduce overfitting [James, 2013; Breiman, 2001]. The treatment effect for each individual in the validation dataset is calculated using the subgroup of individuals in their assigned terminal node based on the interaction tree constructed on the training dataset. Separating the construction of the trees (training dataset) and the estimation of treatment effects (validation dataset) maintains honesty of the random forest [Wager and Athey, 2018]. Individual treatment effects are then averaged across all trees within the random forest. The values of the polygenic response score are the predicted individual treatment effects. A schematic diagram of the random forest method is shown in Figure 2.

**Figure 2:**
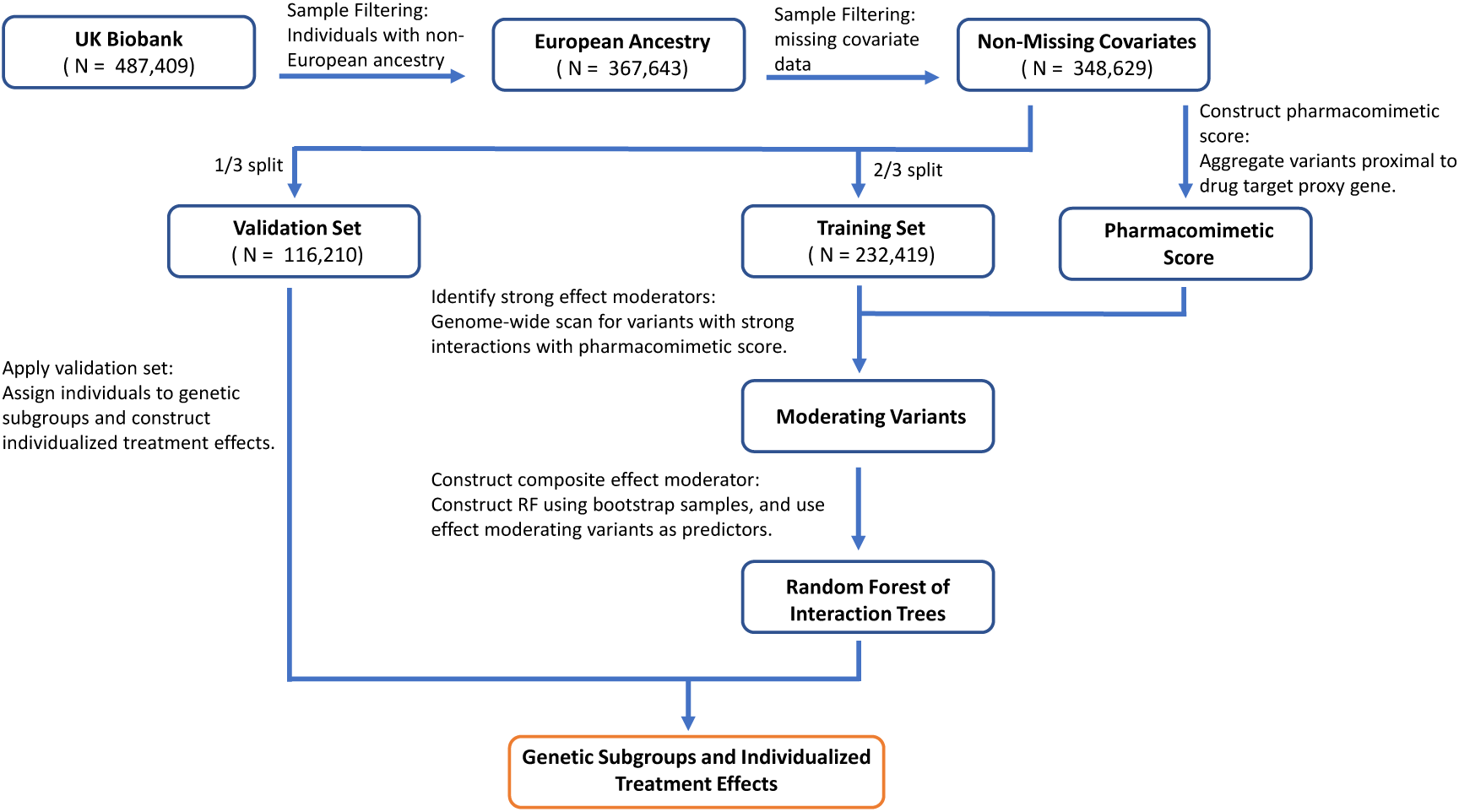
Summary of the random forest of interaction trees method to construct the composite effect modifier (polygenic response score) for the applied example.

### Assessing treatment effect heterogeneity

To assess whether the predicted treatment effects differ by more than expected due to chance alone, we calculated the weighted standard deviation of the predicted treatment effects in the validation set for each tree. We consider an interaction tree with *K* leaf nodes, with the predicted treatment effect for the leaf node subgroups 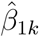 where *k* = 1, …, *K*, and the sample size of the leaf nodes *n*_*k*_ where *k* = 1, …, *K*. The weighted standard deviation for the tree (*σ*) was defined as:

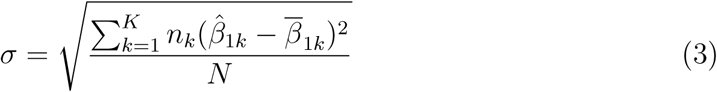

where:

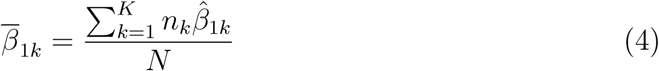

and *N* is the total sample size. We calculated the average of the weighted standard deviations across trees.

We then permuted together the outcomes, covariates, and pharmacomimetic genetic variant (equivalent to randomly assigning individuals to leaf nodes representing genotypic subgroups) for individuals in the validation subset, and calculated the weighted standard deviation of the treatment effects in 1000 permuted validation datasets. If the average weighted standard deviation of the treatment effects is similar in the validation and permuted validation datasets, then there is no more variability in the treatment effect estimates than would be expected by chance alone. We calculated a bootstrap p-value representing the proportion of permuted validation datasets for which the weighted standard deviation is larger for the permuted validation dataset than the original validation dataset.

### Variable importance measure

We calculated variable importance based on the formulation described by Su et al. [2009]. The method involves the permuting each split variable individually, and calculating the decrease in overall interaction when a split variable is permuted. More important split variables should contribute to a higher decrease in overall interaction when permuted, and vice-versa. The variable importance of a variable was then calculated by averaging over its variable importance in each tree of the random forest, separately for the training and validation sets. This allows us to indentify variants that are important effect modifiers.

### Example: effect modification for statins

We applied our method to investigate treatment effect heterogeneity for statins using data from the UK Biobank study. Data were available on 502,682 participants (94% of self-reported European ancestry) recruited between 2006 and 2010 in 22 assessment centres throughout the UK. We considered individual-participant data on 348,629 unrelated individuals of European descent who passed extensive quality control procedures as described in Astle et al. [2016]. Briefly, we excluded participants having non-European ancestry (self-report or judged by genetics), low call rate, or excess heterozygosity (> 3 standard deviations from the mean). We included only one of each set of related participants (third-degree relatives or closer). We also excluded individuals with missing data on LDL-cholesterol, body mass index (BMI), or cholesterol-lowering medication status. LDL-cholesterol was measured on blood serum samples collected at recruitment. For individuals who reported taking cholesterol-lowering medication, the LDL-cholesterol measurement was multiplied by a factor of 1.25 to approximate their LDL-cholesterol level without medication. A pharmacomimetic score was constructed as a weighted score using six genetic variants in or around the *HMGCR* gene region, as previously reported by Ference et al. [2016], and weighting by the associations of the variants with LDL-cholesterol where effect alelles were coded as the LDL-lowering alleles (Web Table A1, Web Figures A1-A2).

In total, 805,426 genetic variants were measured on the UK BiLEVE Axiom array or the UK Biobank Axiom array. Around 40 million further variants were imputed using reference data from the Haplotype Reference Consortium [Bycroft et al., 2018]. We considered all available variants outside of the *HMGCR* gene region (±2 megabase pairs) with a minor allele frequency > 0.05 and an info score > 0.5 as potential moderating variants. Interaction was assessed for each moderating variant in turn using linear regression (equation 1) with main effect terms for the pharmacomimetic score and moderating variant, an interaction term between the pharmacomimetic score and moderating variant, and covariates (age, sex, BMI, and five principal components of ancestry). All variants with a p-value for the interaction term below a given significance threshold were clumped based on correlation (variant removed if *r*^2^ > 0.3 against index variant), with the variant having the lowest p-value for interaction being preferentially selected. The set of independent variants from the clumping procedure were then taken forward to the random forest of interaction trees method. We considered interaction significance thresholds between *p* < 10^−4^ and *p* < 3 × 10^−6^ and minimal node sizes of 5000, 10 000, 20 000, 30 000, and 40 000.

We note that our example differs somewhat from a standard application of Mendelian randomization, in that we do not consider genetic associations with an outcome variable. We restrict our interest to the genetic associations with LDL-cholesterol for two reasons: first, the causal effect of LDL-cholesterol on coronary artery disease risk has been well-established and differences in coronary artery disease risk have been shown to be log-linear in the change in LDL-cholesterol in both trials [Cholesterol Treatment Trialists’ Collaboration, 2010] and Mendelian randomization investigations [Ference et al., 2012]; and second, because interactions in the genetic associations with coronary artery disease are more difficult to detect: coronary artery disease is less proximal to the genetic variants, and the disease variable is binary and relatively uncommon. We assume that any observed heterogeneity in the genetic associations with LDL-cholesterol for the pharmacomimetic variant (and, by inference, in the effect of statins on LDL-cholesterol) would lead to heterogeneity in the genetic associations with coronary artery disease risk (and, by inference, in the effect of statins on coronary artery disease risk).

## Results

### Baseline characteristics of participants

Baseline characteristics of UK Biobank participants in the analytic sample are presented in Table I both for the whole sample, and for individuals with pharmacomimetic score above and below the median value. The pharmacomimetic score was not associated with age, sex, or blood pressure. There was an association of the score with BMI, although the magnitude of association was small. This motivates the inclusion of BMI as a covariate in the interaction tests. There was a strong association of the pharmacomimetic score with LDL-cholesterol and with current use of cholesterol-lowering medication, as expected.

**Table I:**
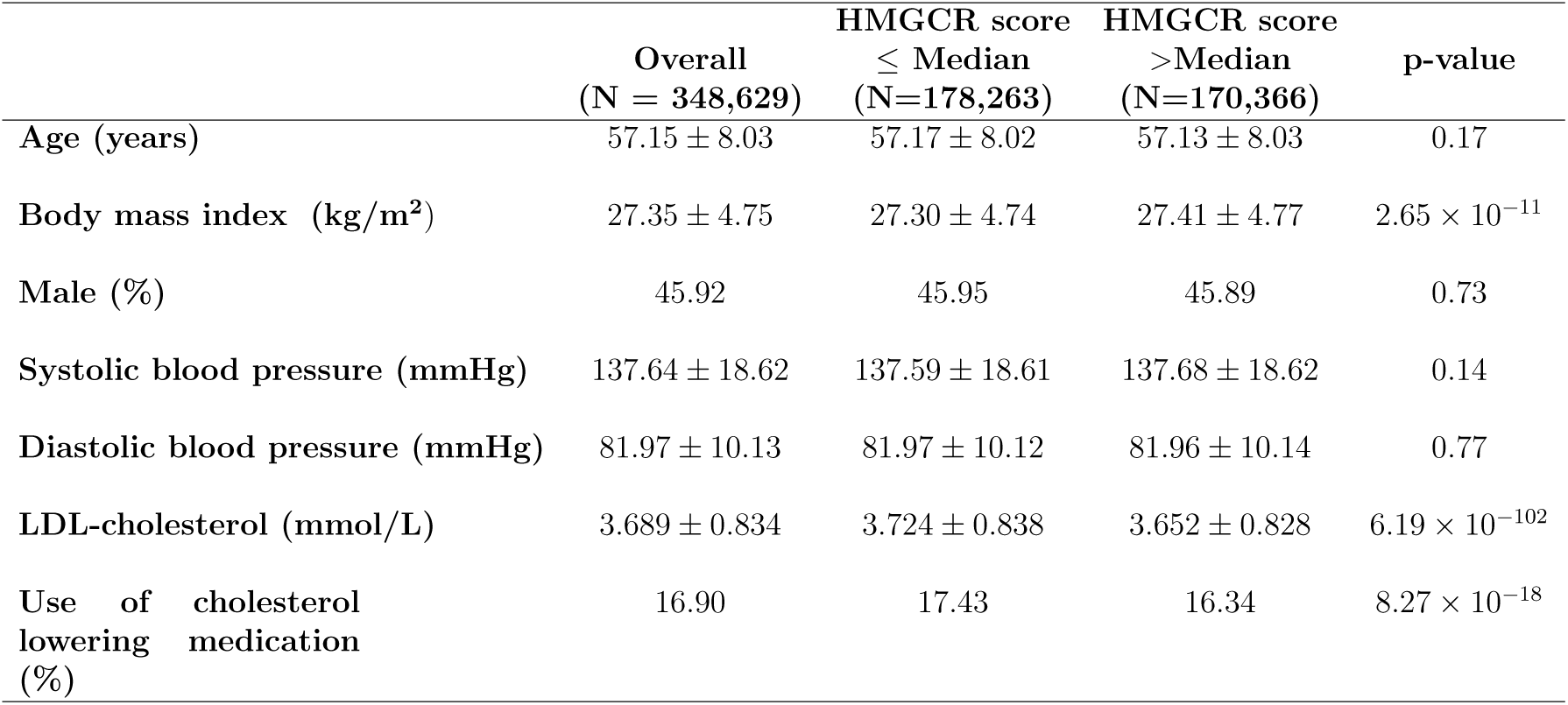
Baseline characteristics of participants: Baseline characteristics (mean and standard deviation, or percentage) of the European ancestry subset of UK Biobank. P-values for differences in characteristics between the subgroups below and above the median HMGCR score are calculated using a t-test for continuous traits and a *χ*^2^ test for categorical traits.

### Gene–gene interactions

No individual genetic variants were found that had a gene–gene interaction at a genome-wide significance level (Web Figure A3). The quantile–quantile plot suggested that there was minimal inflation due to population stratification, and the distribution of interactions was no stronger than would be expected due to chance alone (Web Figure A4).

### Predicted treatment effects

Predicted treatment effects for all individuals in the validation dataset are displayed in Figure 3. These effects can be interpreted as values of the polygenic response score. For more stringent values of the p-value threshold, the distribution of predicted treatment effects is irregular due to the small number of moderating variants, whereas for less stringent values, the distribution approximates a normal distribution. Effect estimates are similar for all individuals in the population at all parameter values for the interaction significance threshold and minimal node size. No individuals had an predicted treatment effect that was positive, or even close to zero. This means that no genetically-defined subgroup of the population was identified that would not be expected to benefit from statin treatment.

**Figure 3:**
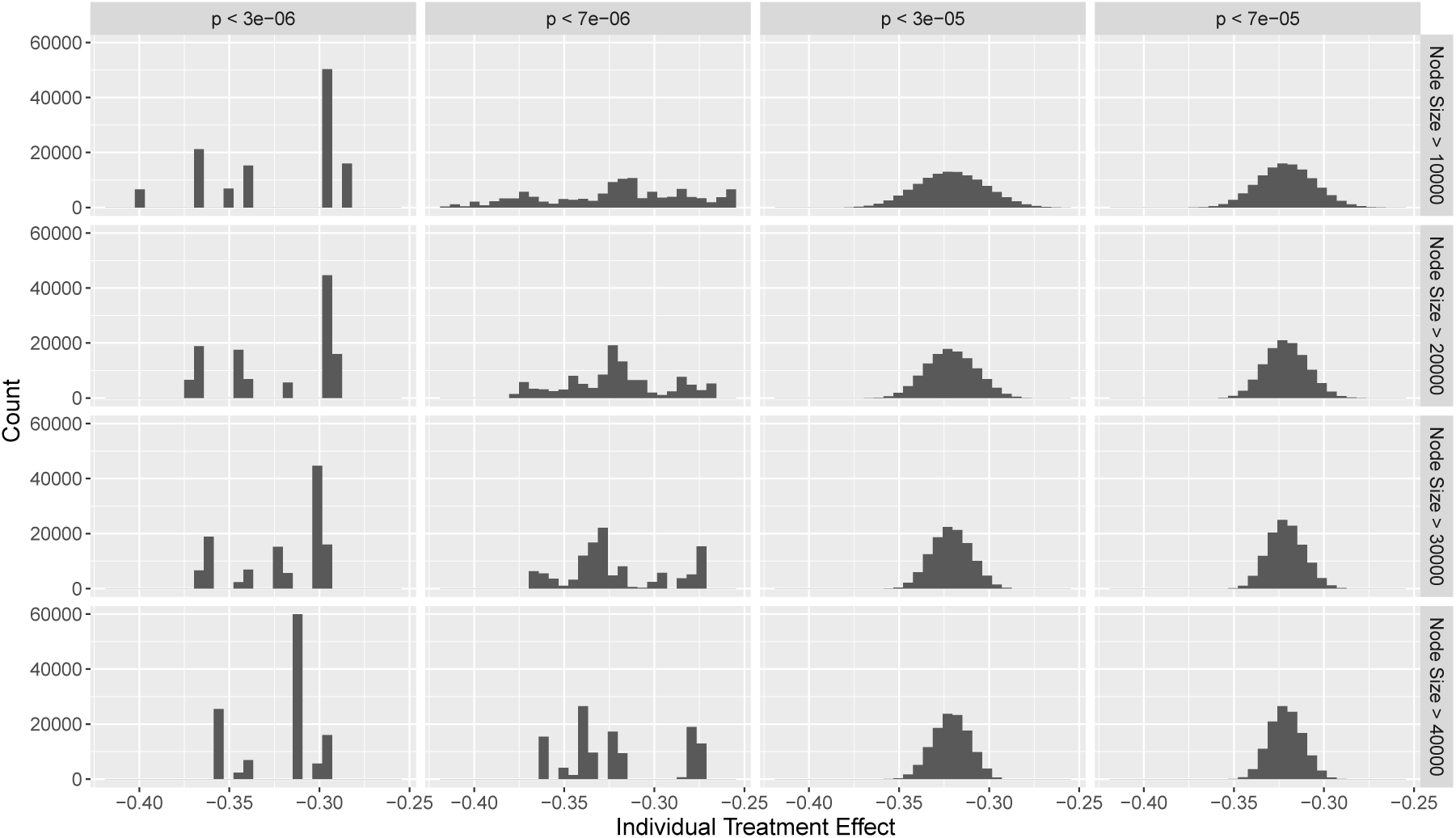
Histograms of predicted individual treatment effects estimated using random forest method with different minimum node sizes and different p-value thresholds for including moderating variants in the analysis.

Figure 4 shows the average difference between the weighted standard deviation of treatment effect estimates in the validation and bootstrap-permuted validation datasets. While the difference was generally positive, indicating higher identified heterogeneity in the validation subset, it was small throughout. The bootstrap p-value was only 0.18 at its minimum value with node size of 40 000 and interaction p-value threshold of 7 × 10^−6^ (Web Figure A6). Hence the predicted treatment effects were no more variable than would be expected due to chance alone.

**Figure 4:**
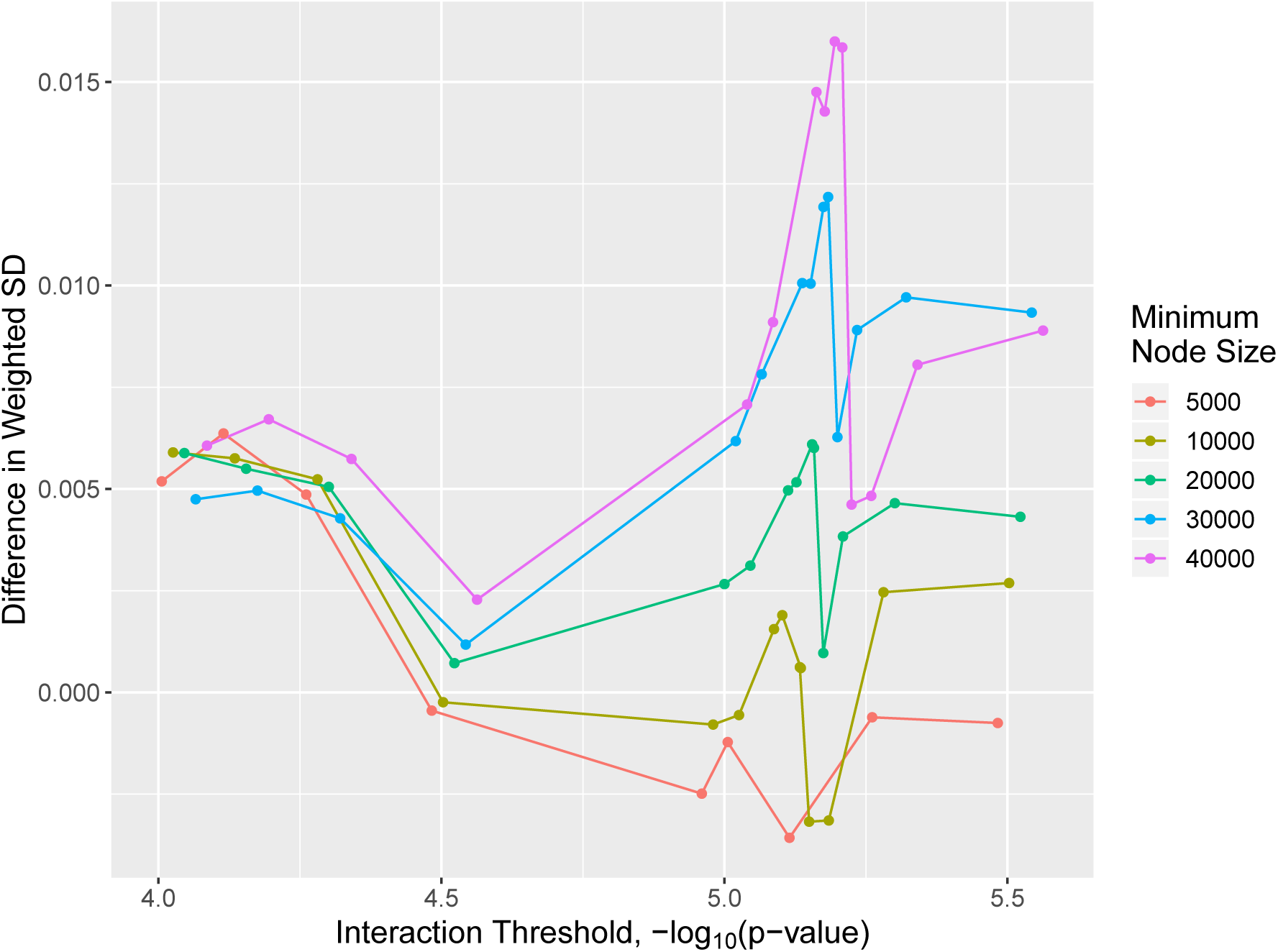
Average difference in weighted standard deviation of the predicted subgroup treatment effect estimates calculated in the validation dataset and boostrap-permuted validation set based on trees estimated in the training dataset. Positive differences indicate higher heterogeneity of predicted subgroup treatment effects identified in true validation dataset compared to the bootstrap-permuted validation set.

### Variable importance measure

Variants having the strongest interactions are displayed in Web Figure A5, together with their variable importance measures. Only two variants (rs162724 and rs12884142) displayed positive variable importance in both the training set and validation datasets, which suggests the generalizability of these effect modifiers.

rs162724 is an intronic variant (minor allele frequency of 0.16 and interaction p-value of 1.2 × 10^−6^) located proximal to the *GRM7* gene on chromosome 3. Previous genome-wide association studies have found strong associations of variants within the *GRM7* gene region with major depressive disorder, schizophrenia, and the efficacy of antipsychotic medication [Need et al., 2009; Stevenson et al., 2016; Shyn et al., 2011; Sacchetti et al., 2017]. The use of some antipsychotics has been found to be associated with altered risk of hyperlipidaemia, and in some studies with elevated LDL-cholesterol levels [Meyer and Koro, 2004; Saari et al., 2004]. Thus, it is possible that the variant (rs162724) acts as a proxy for antipsychotic use in our study, which in turn acts an indirect effect modifier of statins by altering cholesterol levels.

## Discussion

In this paper, we have introduced an agnostic approach to combine genetic variants into a composite effect modifier (a polygenic response score) that divides the population into genetic subgroups which are predicted to respond differently to a particular treatment. This approach relies on the principles of Mendelian randomization that pharmacomimetic genetic variants can be treated as if they have been randomized, and can be used as an unconfounded proxy for the treatment. We have illustrated the approach for the effect of statins on LDL-cholesterol levels. In this example, no more heterogeneity in the predicted treatment effect was detected than would be expected by chance alone, the sign of the predicted treatment effect was the same for all individuals in the population, and there was no subgroup of the population for whom the predicted treatment effect was close to zero. Therefore the clinical impact of this finding is low. However, the approach may have more applicability in other contexts.

A proposed setting where our approach may have clinical utility is for CETP inhibition. While CETP inhibitors have generally failed to demonstrate effectiveness in untargeted clinical trials [Schwartz et al., 2012; Lincoff et al., 2017] (although see [HPS3/TIMI55–REVEAL Collaborative Group, 2017]), there is some evidence for a protective effect of dalcetrapib in a particular genotypic subgroup of the population defined by a variant in the *ADCY9* gene region [Tardif et al., 2015]. Our method could be used to further refine this finding by searching for subgroups based on multiple genetic variants, rather than just considering single variant interactions.

Previous attempts have been made to find genetic variants that predict response to treatment [Lewis et al., 2019], including for statins [Postmus et al., 2014], based on data from clinical trials. Our paper makes two additional methodological contributions to the literature: first, we use the Mendelian randomization paradigm, which allows treatment response to be predicted from cross-sectional data, and secondly, we construct a polygenic response score based on multiple variants from across the genome, rather than just individual variants.

Our results provide some evidence in the wider debate as to whether treatment effect heterogeneity is widespread or uncommon. For example, Senn [2018] argues that treatment effect heterogeneity should not generally be expected, meaning that precision medicine approaches are unlikely to exist for many treatments. More extensive investigations are required to judge whether the degree of effect heterogeneity observed in this paper is typical or not.

Our proposed approach has strengths and also weaknesses. While it would be possible to investigate treatment effect heterogeneity more directly in a trial setting, our approach is able to leverage the large sample sizes available in cross-sectional ‘biobank’ data. Biobank samples are often more representative of the general population than clinical trials, meaning that estimates are obtained in a more relevant target population, particularly if the treatment is for primary prevention. Furthermore, lack of efficacy is the major contributor to failure of phase 3 clincal trials [Fogel, 2018]. Our method allows prior prediction of treatment response, so that trials can be conducted in targeted genetic subgroups. However, there are also potential weaknesses. First, there are many reasons why a statistical interaction may be observed that does not correspond to a biological interaction. For example, it may be that moderating variants increase LDL-cholesterol levels, and that the association of *HMGCR* variants with LDL-cholesterol is simply larger in individuals with greater LDL-cholesterol levels. While this is an example of effect modification, the conclusion that individuals with greater LDL-cholesterol levels would benefit more from LDL-cholesterol lowering is not particularly insightful. Secondly, our approach for identifying moderating variants was relatively simple, and more complex approaches could be considered. For example, we may believe that genetic variants with strong interaction effects are likely to also have strong main effects. Rather than considering the p-value for each interaction term in isolation, we could consider the p-values for the main effect and the interaction effect jointly. Thirdly, the genetic associations with LDL-cholesterol are not particularly strong, with the strongest individual per allele genetic association corresponding to a 0.06 mmol/L change in LDL-cholesterol. In contrast, statins can reduce LDL-cholesterol by around 1 mmol/L. Hence our null result may correspond to a lack of power. However, genetic associations typically represent lifelong changes in the trajectory of a risk factor and so the proportional effect on a disease outcome is generally stronger [Burgess et al., 2012], meaning that genetic interactions may be easier to detect. Also, genetic associations do not suffer from lack of adherence that can attenuate effects in trials. Fourthly, our method relies on the assumption that the pharmacomimetic variants can be treated as proxies for the relevant treatment. In practice, there may be ways in which the genetic variant does not mimic treatment use. For example, we would not be able to detect effect modifiers which are drug metabolizers. Genetic polymorphisms in Cytochrome P450, a drug metabolizing enzyme, has been shown to be strongly associated with response to statins [Lynch and Price, 2007; Canestaro et al., 2014]. It is possible that majority of the effect heterogeneity may be attributed to drug metabolism. Finally, our results could be affected by population factors such as ethnicity. Detecting such heterogeneity would be useful, as it would still identify subgroups of the population that have different treatment response. However, varying a treatment regime based on ethnicity would not generally be regarded as precision medicine, as precision medicine seeks to find differences within populations rather than between populations. We have tried to reduce the impact of ethnicity by restricting our analysis to individuals of European descent and adjusting for genomic principal components. However, we cannot rule out a residual effect of population stratification on our results.

In conclusion, we have demonstrated an agnostic genome-wide approach to create a polygenic response score that explains heterogeneity in the predicted effect of a treatment. While the clinical impact of the example demonstrated here is limited, this approach may be useful to detect individuals with particularly strong or weak predicted response to particular treatments, leading to opportunities for precision medicine.

## Data Availability

The data that support the findings of this study are available on application to any bona fide researcher. See https://www.ukbiobank.ac.uk/ for more details.

## Acknowledgements

The research has been conducted using the UK Biobank Resource.

## Data availability statement

The data that support the findings of this study are available on application to any *bona fine* researcher. See https://www.ukbiobank.ac.uk/ for more details.

## Appendix

**Web Table A1:** SNPs selected for the construction of pharmacomimetic score in or around the *HMGCR* gene region: rsid, p-value (P) and effect size for association with LDL-cholesterol in mmol/L (Beta), standard error (SE), chromosome (Chr) and position number in hg19 (Pos), LDL-lowering coded allele / effect allele (A1), non-effect allele (A2) and effect allele frequency (EAF) calculated from included samples within the UK Biobank.

| rsid | P | Beta | SE | Chr | Pos | A1 | A2 | EAF |
| --- | --- | --- | --- | --- | --- | --- | --- | --- |
| rs12916 | $1 \times 10^{-226}$ | -0.0647 | 0.0020 | 5 | 74656539 | T | C | 0.60 |
| rs17238484 | $2 \times 10^{-114}$ | -0.0535 | 0.0024 | 5 | 74648496 | G | T | 0.77 |
| rs10066707 | $2 \times 10^{-96}$ | -0.0427 | 0.0021 | 5 | 74560579 | G | A | 0.63 |
| rs5909 | $3 \times 10^{-49}$ | -0.0499 | 0.0034 | 5 | 74656175 | G | A | 0.91 |
| rs2006760 | $2 \times 10^{-47}$ | -0.0358 | 0.0025 | 5 | 74562029 | C | G | 0.80 |
| rs2303152 | $8 \times 10^{-23}$ | -0.0323 | 0.0033 | 5 | 74641707 | G | A | 0.90 |

**Web Figure A1:**
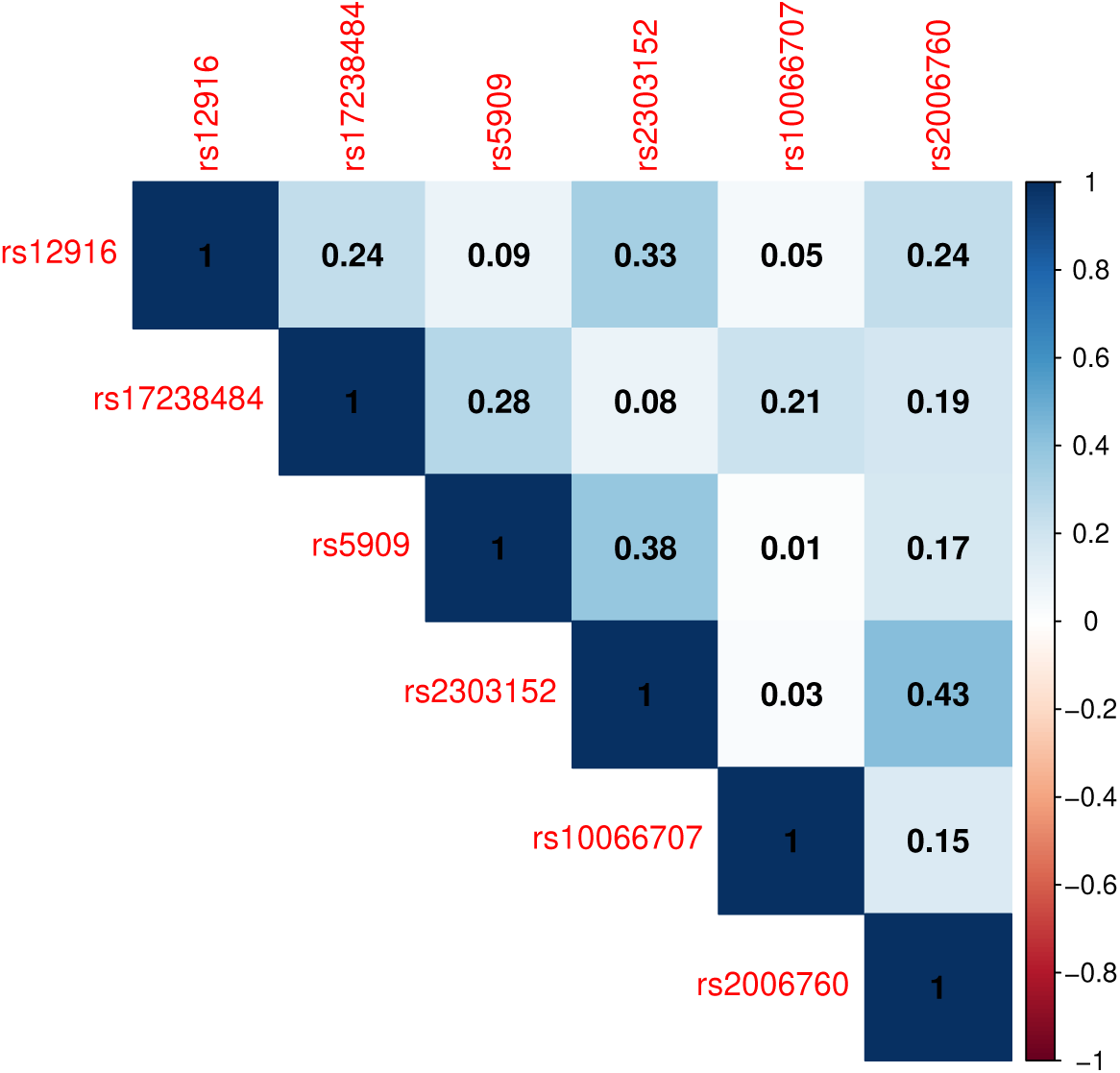
*r*^2^ values illustrating the linkage disequilibrium between SNPs included in the HMGCR pharmacomemimetic score.

**Web Figure A2:**
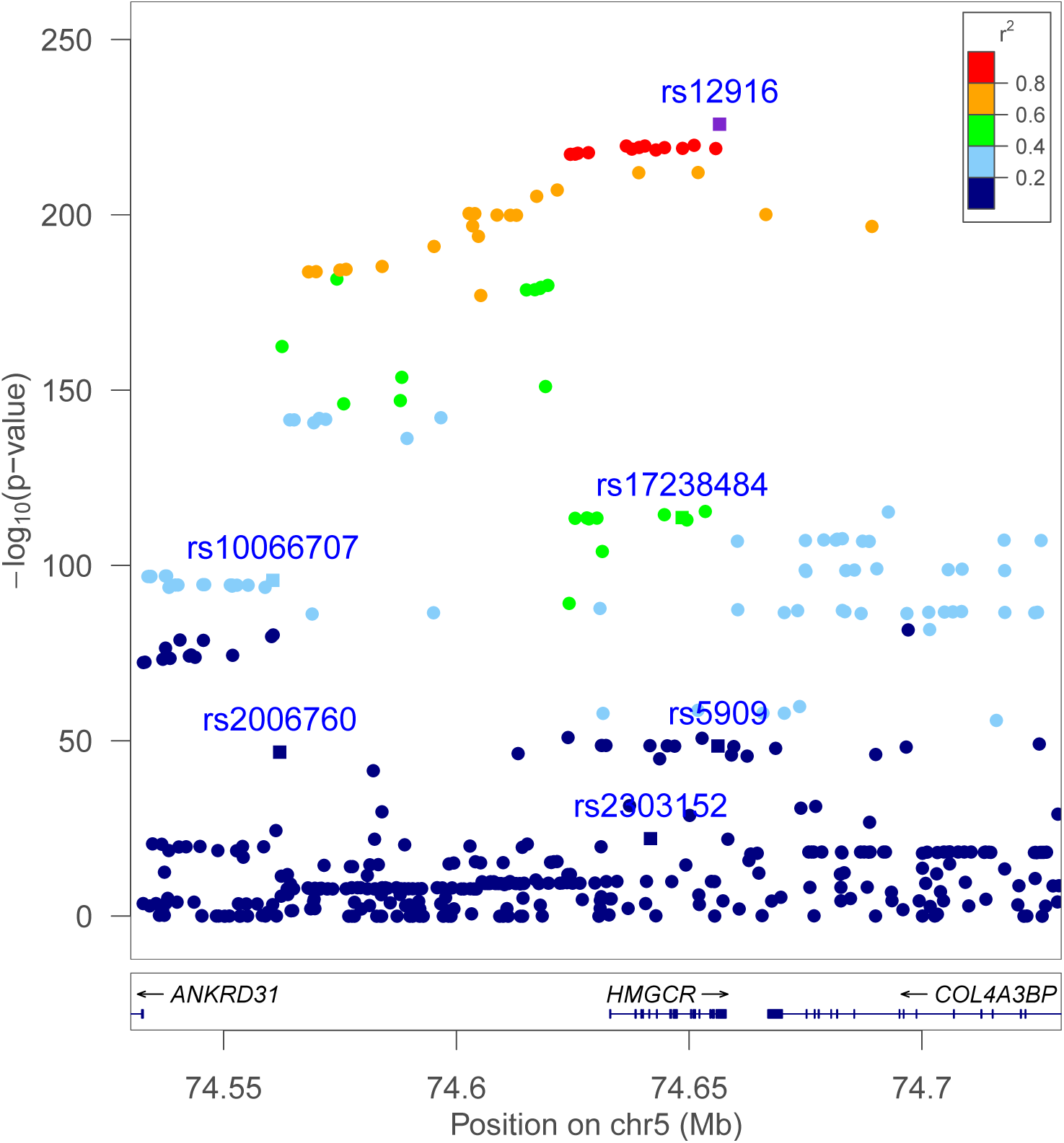
Zoomed-in Manhattan plot of genetic associations with LDL-cholesterol for variants in and around the *HMGCR* gene region.

**Web Figure A3:**
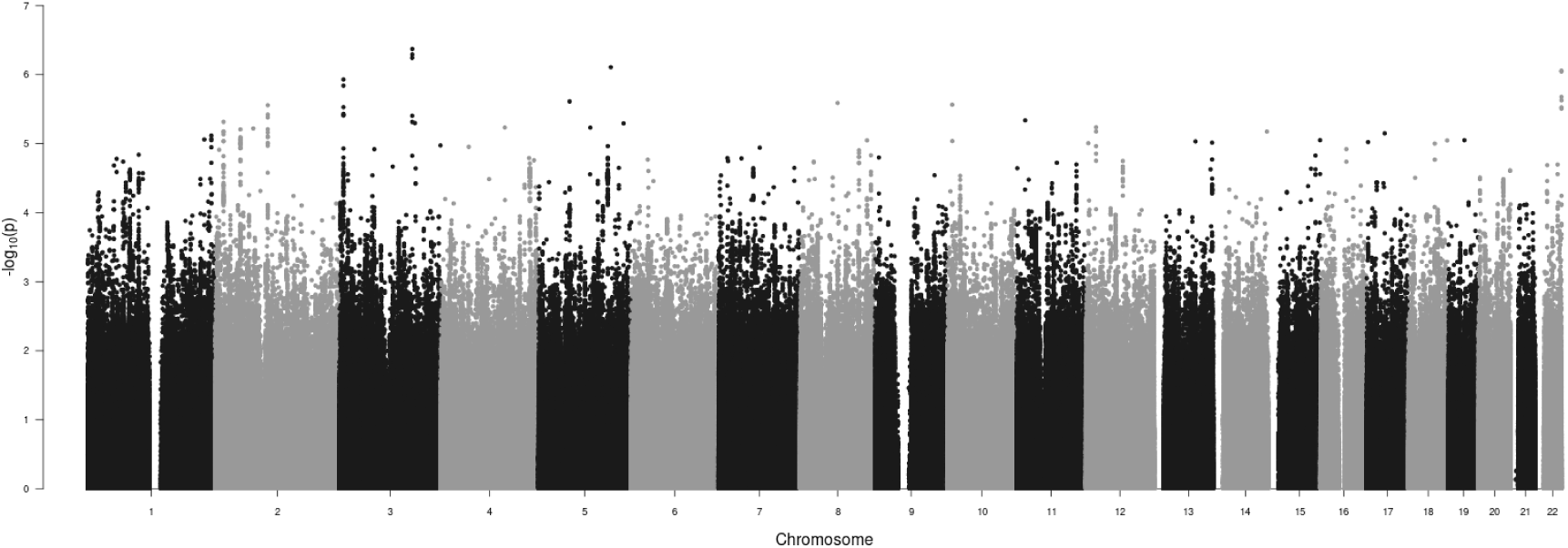
Manhattan plot for interactions, where p-value indicates the strength of interaction between the candidate moderating variant and HMGCR score in their association with LDL-cholesterol.

**Web Figure A4:**
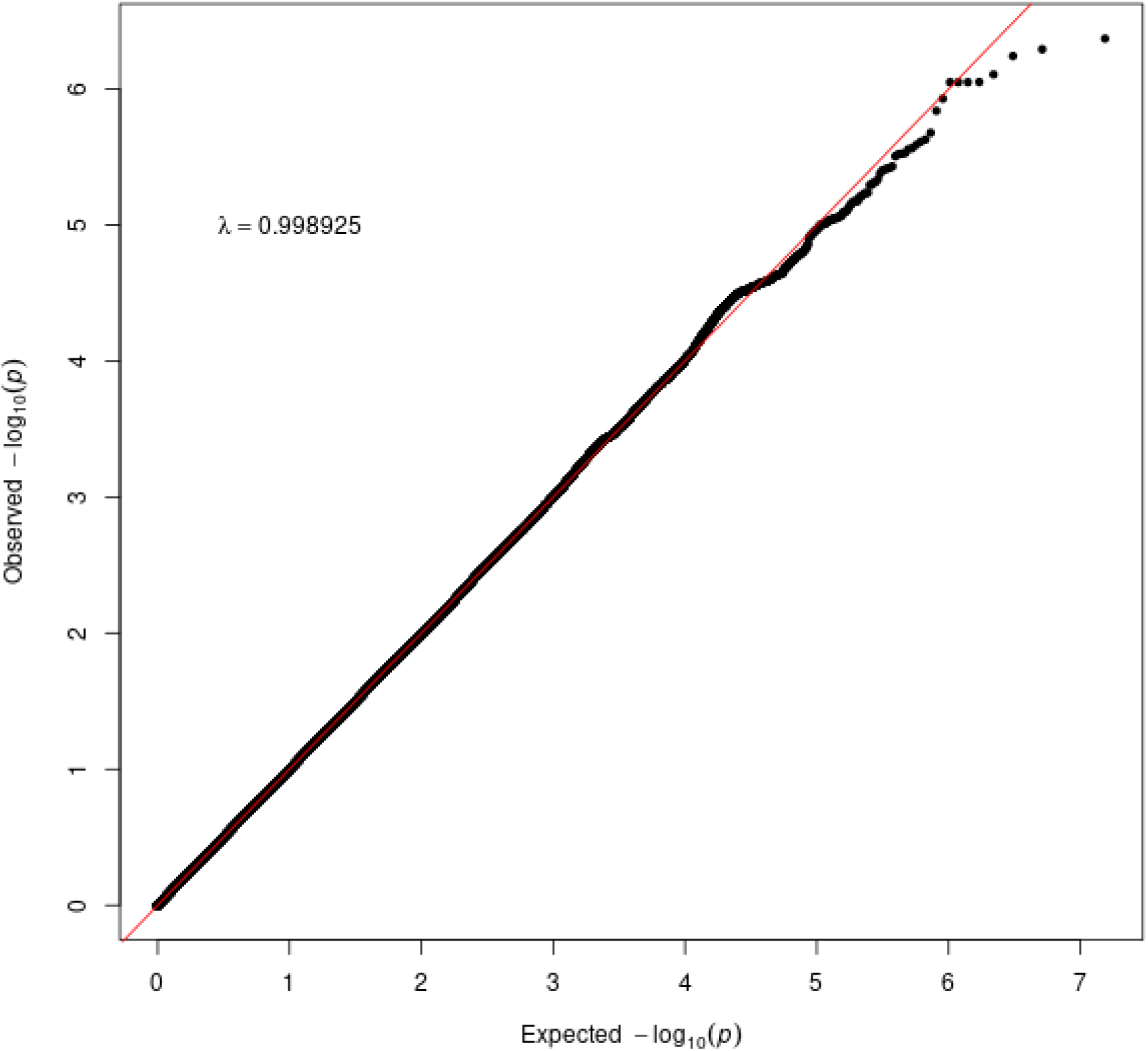
Quantile–quantile plot of p-values for interaction term from linear regression using each candidate moderating genetic variant in turn. The inflation factor *λ* is very close to one, suggesting no inflation due to population stratification.

**Web Figure A5:**
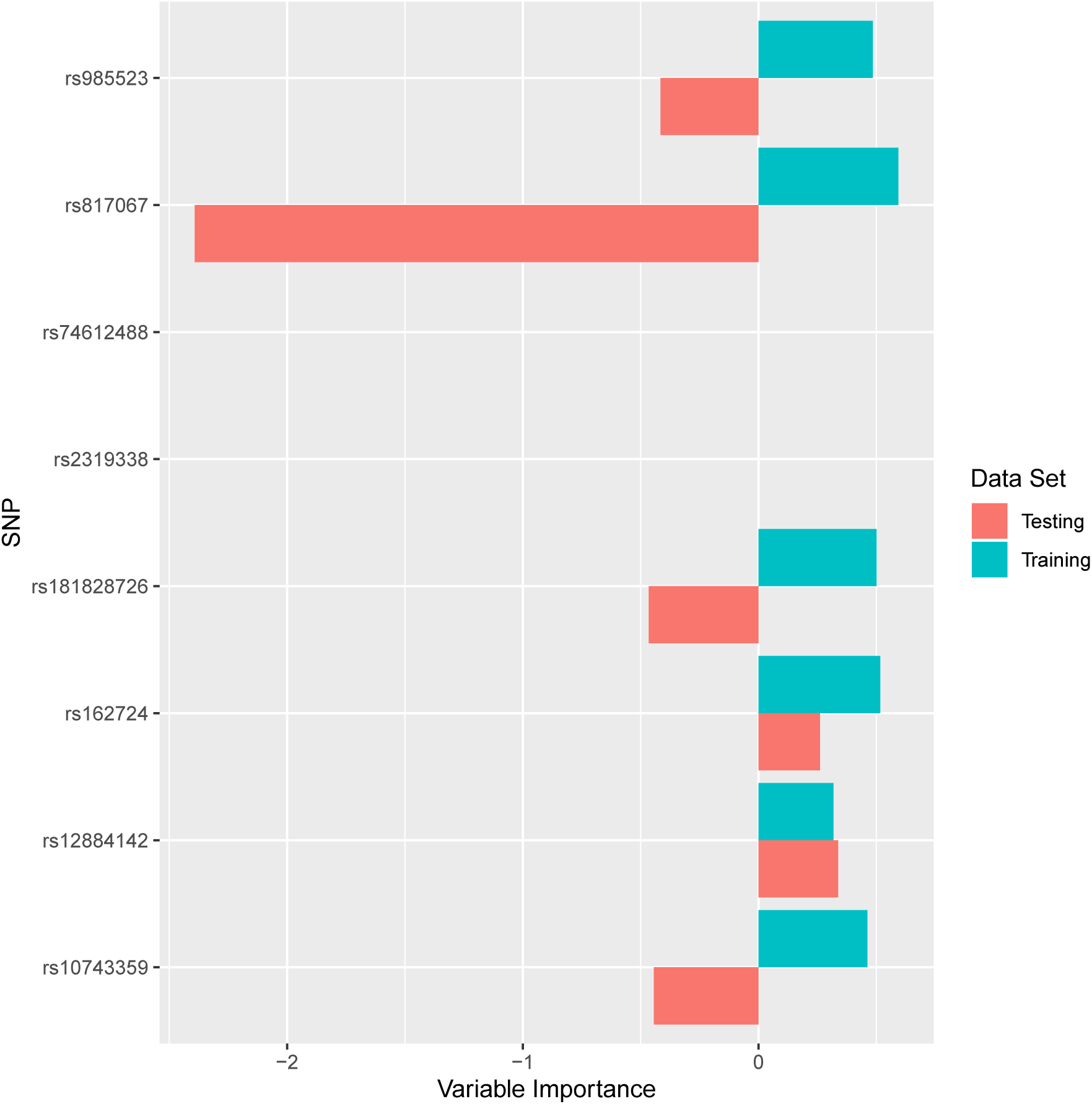
Variable importance of SNPs used in the construction of random forest of interaction trees with significance threshold of *p* < 7 × 10^−6^ and minimum node size of 30 000. In total, 8 SNPs had an interaction p-value below this threshold, although 2 variants were never selected as splitting variants as they divided the validation dataset into subsets that were below the minimum node size.

**Web Figure A6:**
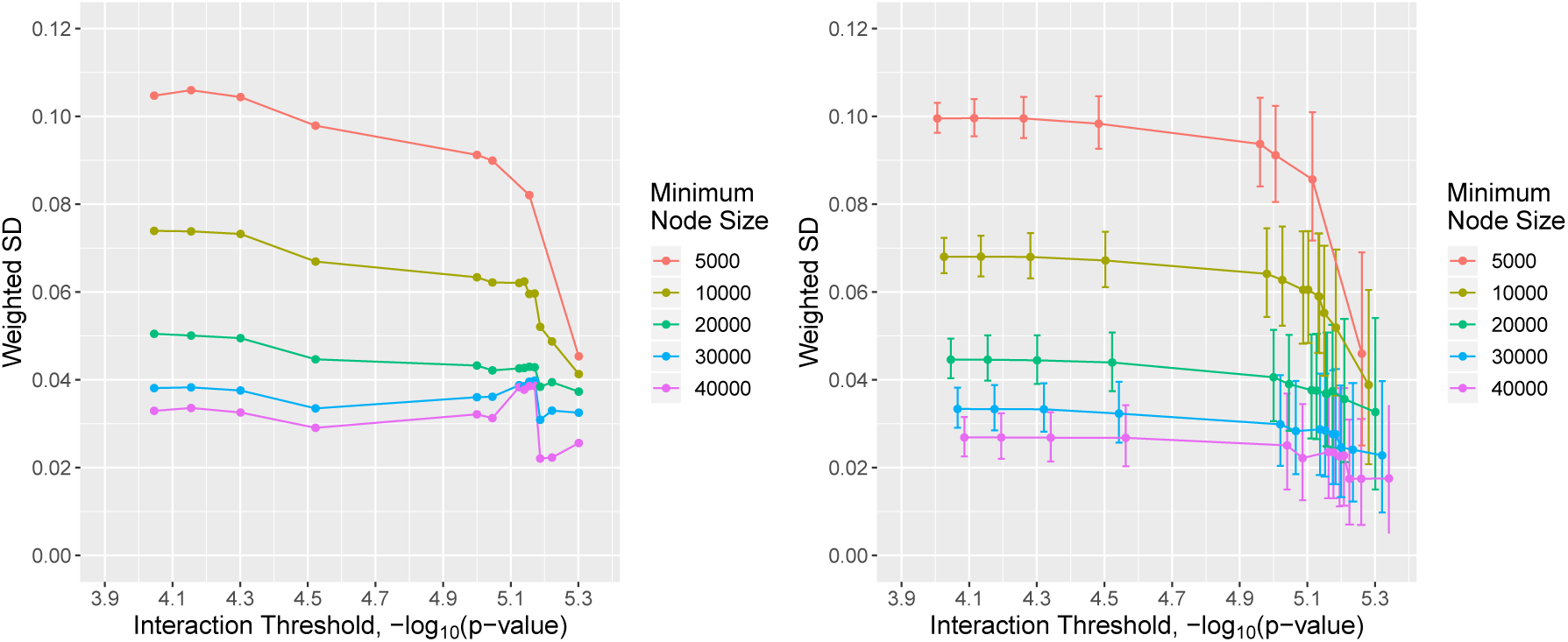
Weighted standard deviation of the predicted individual treatment effect estimates calculated in the validation dataset based on trees estimated in the training dataset for (left) true validation dataset and (right) bootstrap-permuted validation dataset. Error bars indicate the 5th and 95th percentiles of the distribution for the bootstap-permuted datasets.

